# Relationship Between Donor Derived Cell-Free DNA and Tissue-Based Rejection-Related Transcripts In Heart Transplantation

**DOI:** 10.1101/2023.04.07.23288126

**Authors:** Dae Hyun Lee, Ahsan Usmani, Robby Wu, Tammi Wicks, Ryan Burke, Vani Ravichandran, Theresa Wolf-Doty, Ioana Dumitru, Guilherme H. Oliveira, Peter Berman, Benjamin Mackie

**Affiliations:** Heart Failure Center, Heart & Vascular Institute, University of South Florida Morsani College of Medicine, Tampa General Hospital, Tampa, Florida; Heart Transplant Program, Transplant Institute, Tampa General Hospital, Tampa, Florida; Morsani College of Medicine, University of South Florida, Tampa, Florida; CareDx, San Francisco, California

**Keywords:** heart transplant, allograft rejection, donor-derived cell-free DNA, molecular microscopy, gene expression profile, antibody-mediated rejection, acute cellular rejection

## Abstract

**Introduction:** Endomyocardial biopsy (EMB)-based traditional microscopy remains the gold standard for the detection of cardiac allograft rejection, despite its limitation of inherent subjectivity leading to inter-reader variability. Alternative techniques now exist to surveil for allograft injury and classify rejection. Donor-derived cell-free DNA (dd-cfDNA) testing is now a validated blood-based assay used to surveil for allograft injury. The molecular microscope diagnostic system (MMDx) utilizes intragraft rejection-associated transcripts (RATs) to classify allograft rejection and identify injury. The use of dd-cfDNA and MMDx together provides objective molecular insight into allograft injury and rejection. The aim of this study was to measure the diagnostic agreement between dd-cfDNA and MMDx and assess the relationship between dd-cfDNA and MMDx-derived RATs which may provide further insight into the pathophysiology of allograft rejection and injury.

**Methods:** This is a retrospective observational study of 186 endomyocardial biopsy (EMB) evaluated with traditional microscopy and MMDx. All samples were paired with dd-cfDNA from peripheral blood prior to EMB (up to 1 month). Diagnostic agreement between traditional microscopy, MMDx, and dd-cfDNA (threshold of 0.20%) were compared for assessment of allograft injury. In addition, the relationship between dd-cfDNA and individual RAT expression levels from MMDx was evaluated.

**Results:** MMDx characterized allograft tissue as no rejection (NR) (64.5%), antibody-mediated rejection (ABMR) (25.8%), T-cell-mediated rejection (TCMR) (4.8%), and mixed ABMR/ TCMR (4.8%). For the diagnosis of any type of rejection (TCMR, ABMR, and mixed rejection), there was substantial agreement between MMDx and dd-cfDNA (74.7% agreement). All transcript clusters (group of gene sets designated by MMDx) and individual transcripts considered abnormal from MMDx had significantly elevated dd-cfDNA. In addition, a positive correlation between dd-cfDNA levels and certain MMDx-derived RATs was observed. Tissue transcript clusters correlated with dd-cfDNA scores, including *DSAST, GRIT, HT1, QCMAT and S4*. For individual transcripts, tissue *ROBO4* was significantly correlated with dd-cfDNA in both non-rejection and rejection as assessed by MMDx.

**Conclusion:** Collectively, we have shown substantial diagnostic agreement between dd-cfDNA and MMDx. Furthermore, based on the findings presented, we postulate a common pathway between the release of dd-cfDNA and *ROBO4* (a vascular endothelial-specific gene that stabilizes the vasculature) in the setting of AMR, which may provide a mechanistic rationale for observed elevations in dd-cfDNA in AMR, compared to ACR.

## Introduction

The current gold standard for diagnosis of cardiac allograft injury and rejection after heart transplantation is histologic assessment via endomyocardial biopsy (EMB) (1). Diagnostic criteria for acute cellular rejection (ACR) and antibody-mediated rejection (AMR) require the presence of histological features as defined by the ISHLT grading system (2). However, EMB is invasive in nature, has a major complication rate of ∼1%, and traditional histologic interpretation is subject to moderate inter-reader variability (3).

Noninvasive rejection surveillance using donor-derived cell-free DNA (dd-cfDNA) quantification has robust negative predictive value to rule out clinically significant rejection when normal. In the D-OAR study, significant elevations in blood dd-cfDNA levels were associated with acute rejection (4). More recently, dd-cfDNA has been investigated in clinical scenarios to reduce the frequency of EMB. For example, dd-cfDNA assessment 28 days after transplant had a negative predictive value of 99% at a minimum threshold of 0.25% dd-cfDNA, and was thought to safely reduce the need for surveillance biopsy in up to 81% (5, 6). The molecular microscope diagnostics system (MMDx) was developed to quantify intragraft rejection-associated transcripts (RATs) and inform on the probability of rejection without reliance on the histology (7, 8). In prior studies, MMDx results were comparable to EMB-based traditional microscopy but provided additional objective mechanistic insight into the probability of allograft rejection or injury (9).

There is limited literature investigating the relationship between dd-cfDNA and MMDx, particularly in heart transplantation (10-12). Recently, a single-center study showed moderate agreement between MMDx and dd-cfDNA (10). In another study, when dd-cfDNA was used in conjunction with tissue RATs in kidney transplant patients, specificity to rule-in rejection was improved (12). More recently, the Trifecta kidney study showed a strong relationship between dd-cfDNA and expression of RATs associated with AMR, especially with NK cell transcripts and IFN-γ inducible transcripts (11). The aim of this study was to assess the diagnostic agreement between dd-cfDNA and MMDx, and to assess the relationship between dd-cfDNA and MMDx-derived RATs which may provide further insight into the pathophysiology of allograft rejection and injury.

## Methods

### Study Design

This was a retrospective observational cohort study in heart transplant patients that underwent EMB and dd-cfDNA between August 01, 2021 to May 01, 2022 at Tampa General Hospital. Heart transplant patients monitored with dd-cfDNA (AlloSure®, CareDx Inc.; Brisbane CA) within 1 month of traditional EMB and concurrent molecular microscopy surveillance (Molecular Microscopy Diagnostic System; MMDx, One Lambda) were evaluated. University of South Florida Institutional Review Board approved our study as exempt from review (STUDY003322). The current study is in compliance with the International Society of Heart and Lung Transplantation (ISHLT) Ethics Statement. There was no informed consent from subjects due to the retrospective nature of the study.

### Diagnostic Tests

Venous blood for dd-cfDNA was collected through phlebotomy and sent to CareDx, Inc (Brisbane, CA) per standardized protocol. All dd-cfDNA values less than 0.12% were considered as 0.12% in our analysis. Indication for EMB was either routine or for-cause rejection surveillance. Clinical suspicion for rejection was defined as the presence of heart failure symptoms, angina, abnormal hemodynamics, new-onset arrhythmias, abnormal graft function on echocardiography, elevation of dd-cfDNA, or the presence of new coronary allograft vasculopathy. EMB-based histology was graded by the institution’s board-certified pathologists according to the ISHLT’s revised classification for antibody- and cell-mediated rejection (AMR and ACR, respectively) (13). Allograft rejection was categorized by EMB histology as ACR (≥ 2R), AMR (pAMR≥1), and mixed rejection (≥2R and pAMR ≥1). For MMDx, 1-2 EMB samples were sent to One Lambda per standardized protocol. The MMDx results included categorization as No-Rejection (NR), Early-Injury (EI), T-cell mediated rejection (TCMR), antibody-mediated rejection (ABMR), mixed, and possible rejection (possible TCMR, possible ABMR). In addition, the clinically available MMDx reports provided individual RAT expression levels along with further categorization of each transcript as normal, slightly abnormal, or abnormal based on the transcript levels (7). Some of the individual RATs are individual genes or gene clusters (group of genes with common mechanistic pathway, designated by MMDx). In some of our analyses, we used the term “all rejection,” which designates all types of rejection combined into one group, including AMR, ACR, and mixed AMR/ACR. Of note, MMDx uses the terms “TCMR” and “ABMR” to refer to the molecular signatures of histopathologic ACR and AMR, respectively. Thus, throughout the course of this manuscript, the terms “TCMR” and “ABMR” refer to acute cellular and antibody-mediated rejection as assessed by MMDx, while the terms “ACR” and “AMR” refer to histopathologic diagnoses.

### Statistical Analysis

Study data were collected and managed using REDCap electronic data capture tools hosted at the University of South Florida (14). Statistical analysis was performed through GraphPad Prism 9 (GraphPad Software, LLC) and RStudio (15). Data were presented as number/ percentage (for categorical variables) or as mean with standard deviation (mean ± SD) / median with interquartile range (median [IQR]), depending on the normality of the data. Percent agreement and Cohen Kappa coefficients were assessed between the three diagnostic modalities (dd-cfDNA, MMDx, and histology). Percent agreement is classified into low (0-40%), moderate (40-60%), substantial (60-80%), or near-perfect (80-99%) agreement (16). Analysis of variance (ANOVA) with post-hoc Kruskal-Wallis nonparametric tests was performed for comparison between groups. Spearman correlation analysis was performed to investigate the relationship between dd-cfDNA and individual RAT expression from MMDx. Receiver operating characteristic (ROC) analysis was performed for dd-cfDNA (the threshold for normal of <0.20%) and allograft rejection assessment by MMDx or traditional microscopy. A P-value of less than 0.05 (p<0.05) was considered statistically significant. All statistical analyses were performed through R Studio (15). Some graphs (ROC and bar graphs) were generated using GraphPad Prism.

## Results

A total of 186 paired dd-cfDNA, histology, and MMDx samples from 134 patients were analyzed. Baseline patient characteristics are summarized in **Table 1**. The median time between transplant and EMB was 1473 days (IQR 465-2985 days). The indication for EMB was routine surveillance (62.4%; n=116) and for-cause rejection surveillance (37.6%; n=70). The presence of rejection by histology versus MMDx, respectively, are as follows: no rejection (69.4% [n=129] vs. 64.5% [n=120]), antibody-mediated rejection (27.4% [n=51] vs. 25.8% [n=48]), cellular-mediated rejection (1.6% [n=3] vs. 4.8% [n=9]), and mixed (1.6% [n=3] vs. 4.8% [n=9]).

**Table 1.** Baseline Patient Characteristics

| **Variable** | **Total N= 134** |
| --- | --- |
| Age | 56.0 ± 14.0 |
| Sex |  |
| Male | 93 (69.4%) |
| Female | 41 (30.6%) |
| Ethnicity |  |
| Caucasian | 80 (59.7%) |
| Black | 25 (18.7%) |
| Hispanic | 21 (15.7%) |
| Asian | 3 (2.2%) |
| Other | 5 (3.7%) |
| Etiology of Heart Failure |  |
| Nonischemic Cardiomyopathy | 82 (61.2%) |
| Ischemic Cardiomyopathy | 47 (35.1%) |
| Congenital | 5 (3.7%) |

Abbreviations: AMR, Antibody-mediated rejection; ACR: acute cellular rejection.
Data presented as mean ± standard deviation for age, and number (percentage of the whole) for the rest of the variables.

### Concordance analysis: MMDx and dd-cfDNA

Concordance analyses between the three diagnostic modalities (MMDx, traditional microscopy, and dd-cfDNA) are summarized in **Table 2**. The percent agreement between MMDx and traditional microscopy for any type of rejection was substantial (75.8%). The percent agreement between MMDx and traditional microscopy in AMR/ACR were both high (76.9% and 91.4%, respectively). The percent agreement between MMDx and dd-cfDNA (threshold γ0.20%) for any type of rejection was substantial (74.7%). The percent agreement between MMDx and dd-cfDNA was higher in ABMR, compared to TCMR (75.3% vs. 54.3%, respectively). Finally, the percent agreement between traditional microscopy and dd-cfDNA for any type of rejection was 67.7%, similar to the percent agreement between MMDx and dd-cfDNA.

**Table 2.** Diagnostic modality comparison with each rejection type.

| **Diagnostic Modality** | **Rejection Type** | **Percent Agreement** | **Cohen Kappa Coefficient** | **P Value** |
| --- | --- | --- | --- | --- |
| MMDx and traditional microscopy | ABMR/AMR | 76.9 | 0.448 | <0.001 |
|  | TCMR/ACR | 91.4 | 0.299 | <0.001 |
|  | Any Rejection | 75.8 | 0.455 | <0.001 |
| MMDx and dd-cfDNA | ABMR | 75.3 | 0.505 | <0.001 |
|  | TCMR | 54.3 | 0.086 | 0.0472 |
|  | Any Rejection | 74.7 | 0.495 | <0.001 |
| EMB and dd-cfDNA | AMR | 66.1 | 0.323 | <0.001 |
|  | ACR | 53.2 | 0.0645 | 0.0128 |
|  | Any Rejection | 67.7 | 0.355 | <0.001 |

Abbreviations: MMDx: molecular microscopy diagnostic system; dd-cfDNA: donor-derived cell-free DNA; EMB: endomyocardial biopsy; AMR: antibody-mediated rejection; ACR: acute cellular rejection

Next, we compared the median level of dd-cfDNA in various types of rejection categories by MMDx (No Rejection or NR, ABMR, TCMR, and mixed rejection; **Figure 1A**). The median dd-cfDNA in NR was 0.10% (IQR: 0.1-0.3%). The median dd-cfDNA levels in ABMR, TCMR, and mixed rejection were 1.1% (IQR 0.4-2.0%), 0.20% (IQR 0.2-0.6), and 1.9% (IQR 1.4-2.3%), respectively. ABMR and mixed rejection, as assessed by MMDx, had higher levels of dd-cfDNA compared to the NR group (p<0.001). However, TCMR was not different compared to NR (p=0.238). In addition, in the absence of any rejection (N=120), the median level of dd-cfDNA was higher in the presence of injury (by MMDx) (0.2 [IQR: 0.1-0.6]; N=13), compared to the absence of injury (0.1 [IQR: 0.1-0.2]; N=107; p=0.110), although not statistically significant. At a threshold of dd-cfDNA level of 0.2%, the area under the curve (AUC) for the combined use of MMDx and dd-cfDNA in predicting rejection was 0.73 for TCMR, 0.86 for ABMR, and 0.84 for any type of rejection (**Figure 1B-D**).

**Figure 1.**
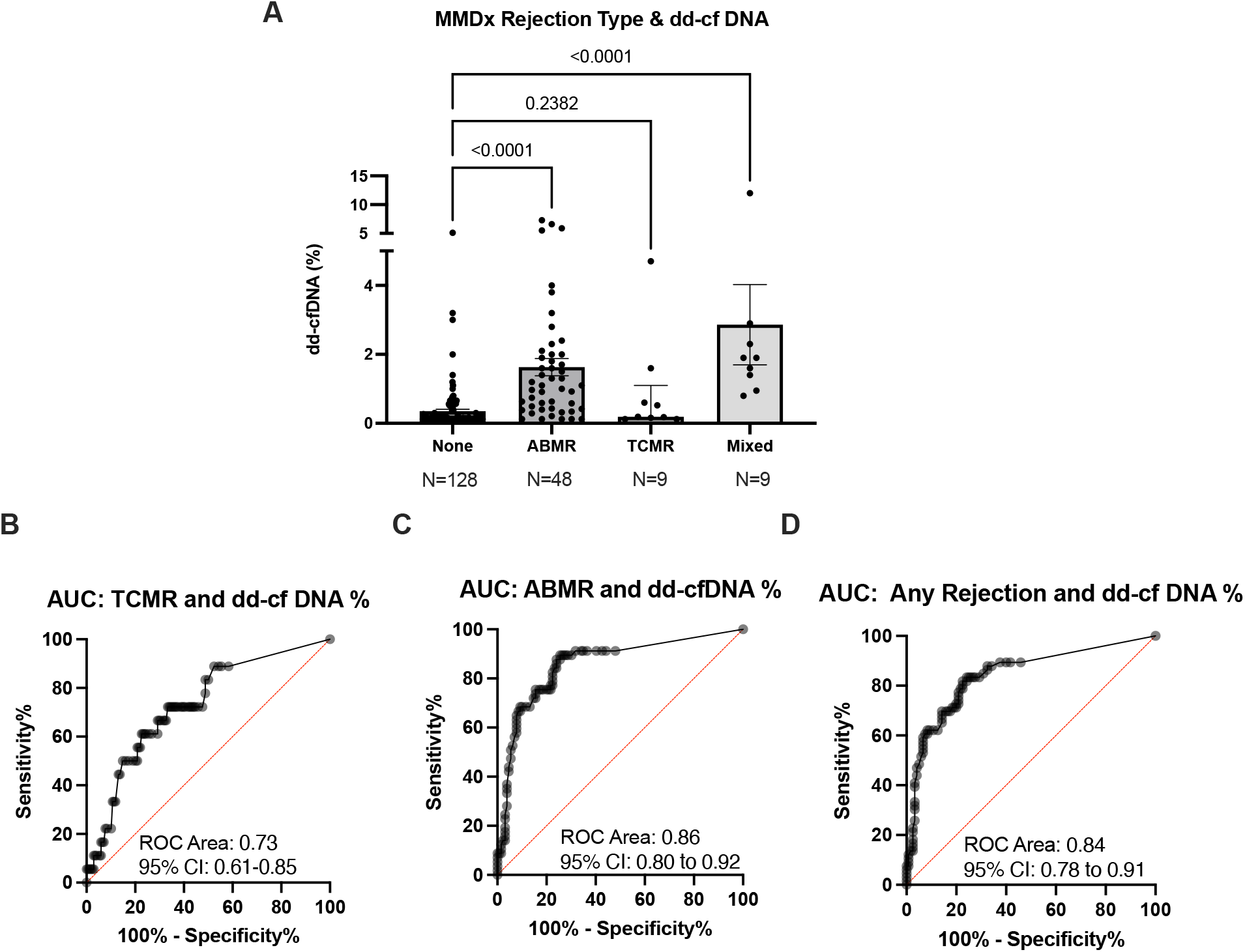
Relationship between MMDx and dd-cfDNA. **A**, Bar graph of dd-cfDNA (%) in different types of rejections as assessed by MMDx (median with interquartile range). Comparison between groups was performed through Kruskal-Wallis test with multiple comparisons. Antibody-mediated rejection (ABMR) and mixed rejection (ABMR and TCMR) had higher levels of dd-cfDNA compared to no rejection. T Cell-mediated rejection (TCMR) did not have different levels of dd-cfDNA compared to no rejection. **B-D**, Area under the curve (AUC) for different types of rejections (ABMR, TCMR, or Any) and dd-cfDNA% at a cut-off of 0.20%. Abbreviations: MMDx: molecular microscopy; dd-cfDNA: donor-derived cell-free DNA. ABMR: antibody-mediated rejection; TCMR: T-cell mediated rejection; ROC: Receiver operating characteristics; AUC: area under the curve

### MMDx-based Rejection-Associated Transcript and dd-cfDNA

Next, we investigated the relationship between the MMDx-derived tissue RATs and dd-cfDNA **(Figure 2 and Table 3)**. In all fifteen RAT classifications, samples defined as slightly abnormal or abnormal had higher levels of dd-cfDNA when compared to those defined as normal (**Table 3)**. Expression level of the 15 individual RAT and dd-cfDNA levels are shown in the scatterplot in **Figure 2**. *DSAST* (endothelial activation), *GRIT* (IFN-γ response), *NKB* (NK-cell burden), *QCMAT* (macrophage burden), *HT1* (parenchymal injury), *IFNG* (IFN-γ), *QCAT* (Cytotoxic T-cell infiltration) and *ROBO4* (endothelial senescence) moderately correlated with dd-cfDNA in Spearman correlation (coefficient of greater than 0.5). Of these, the only RAT that was an individual transcript, rather than a transcript cluster, was *ROBO4* (R=0.53; p<0.001).

**Table 3.**
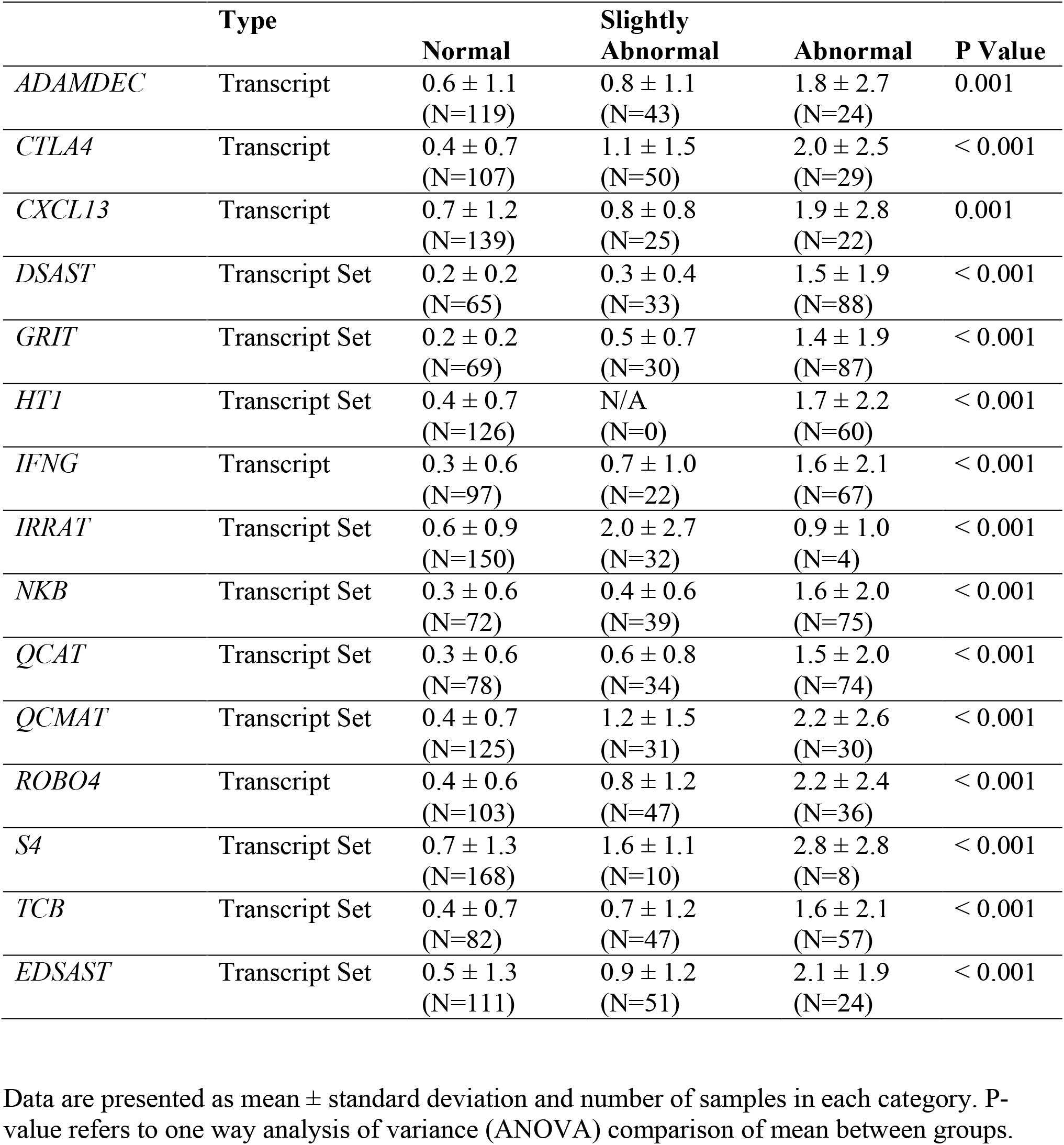
Average level of dd-cfDNA based on the categorization of transcript expression by MMDx

**Figure 2.**
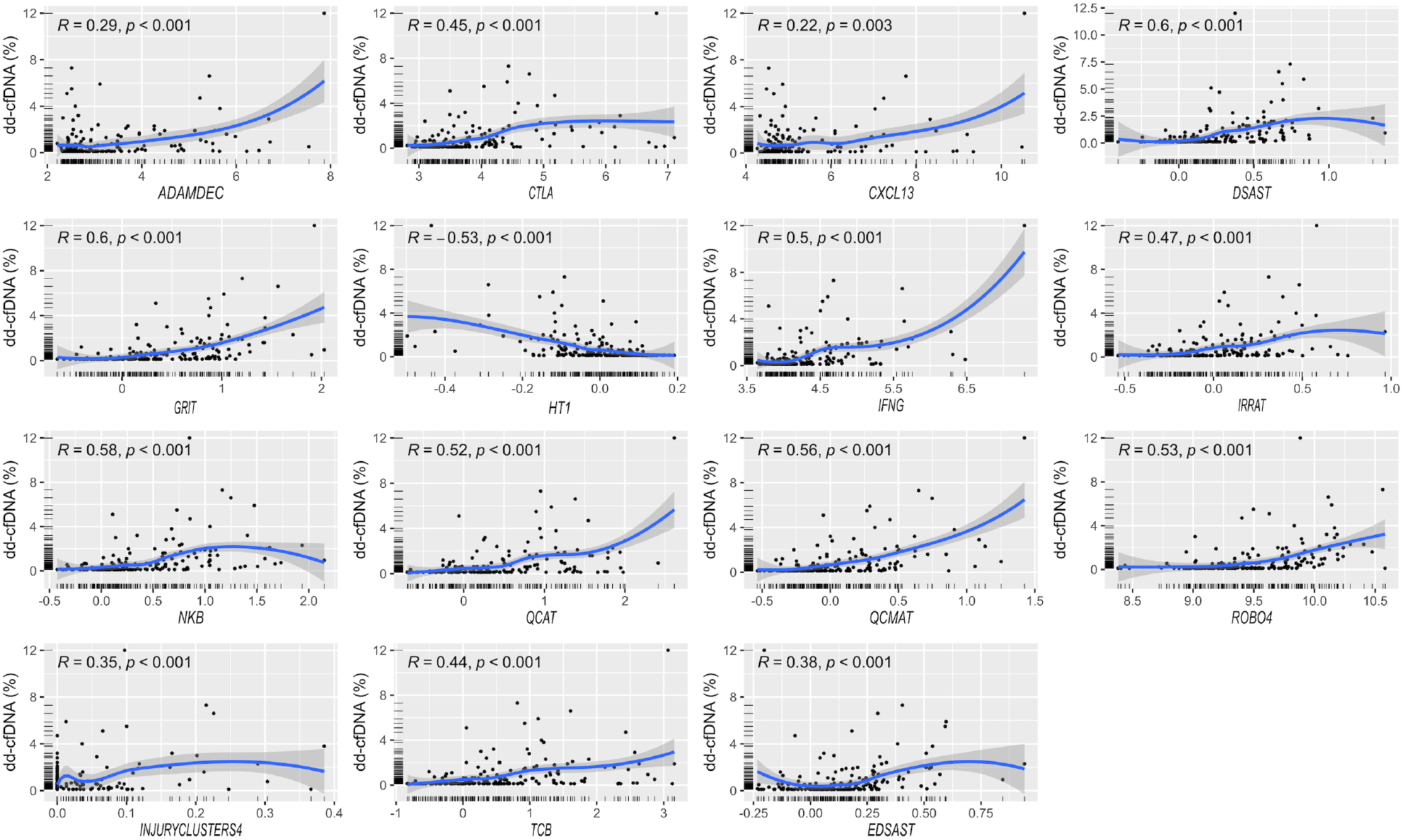
MMDx-based tissue rejection-associated transcripts (RATs) expression profile and dd-cfDNA levels. Scatterplots quantifying % dd-cfDNA as a function of expression of individual RATs within the MMDx signature. The dark blue line indicates the best regression line-of-fit between the RAT expression levels and dd-cfDNA. The dark grey area indicates the 95% confidence interval of the best regression line-of-fit. Abbreviations: MMDx: molecular microscopy; dd-cfDNA: donor-derived cell-free DNA.

Next, we investigated the relationship between RAT expression level and dd-cfDNA based on the presence or absence of rejection by MMDx (**Figure 3**). In both the rejection and no-rejection groups, several RAT’s expression levels had statistically significant correlations with dd-cfDNA levels (*DSAST, GRIT, HT1, QCMAT, ROBO4, and INJURYCLUSTER/S4)*.

**Figure 3.**
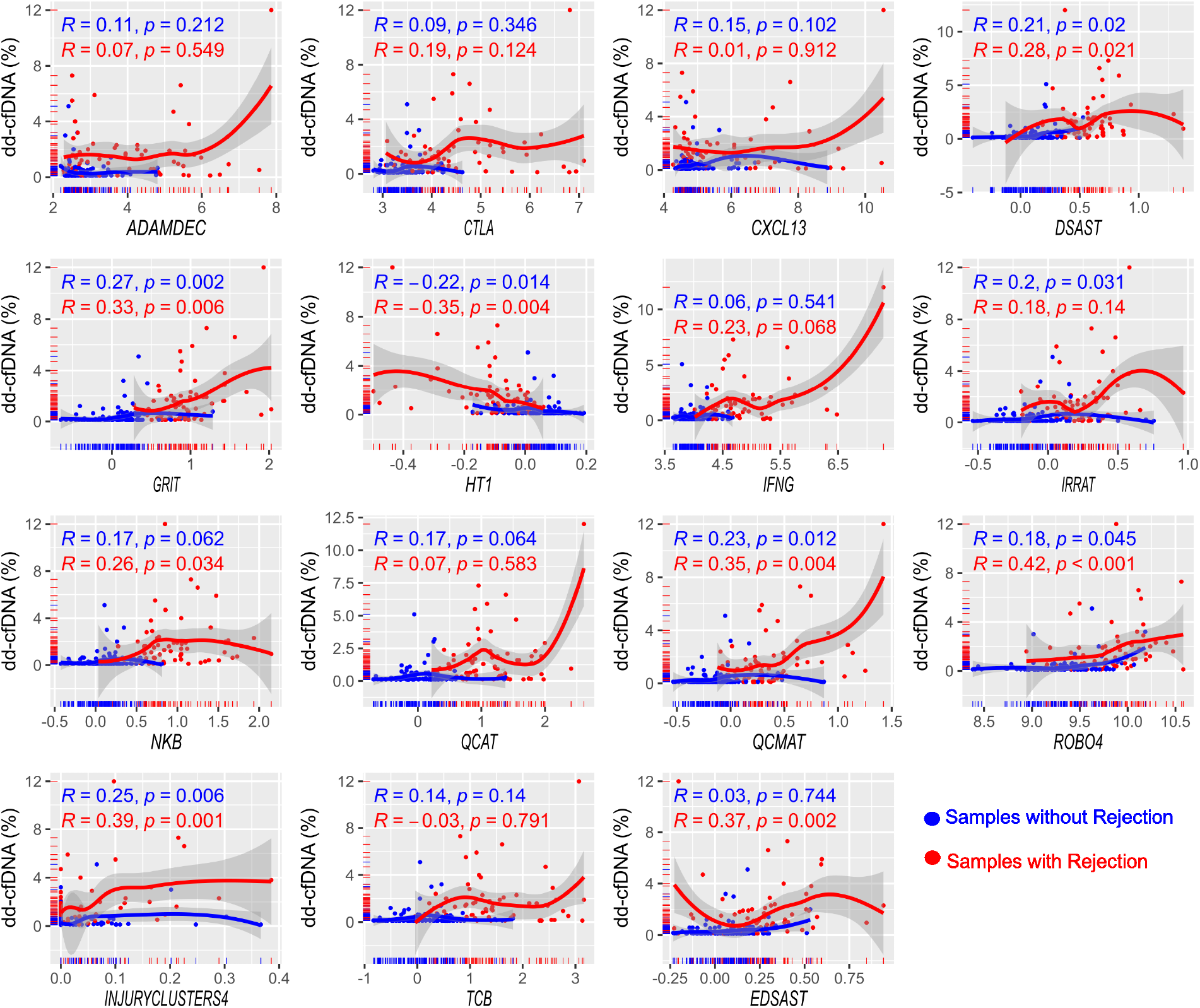
dd-cfDNA level and MMDx-based rejection-associated transcripts in rejection and no rejection patients. The red dots/regression line represents the rejection group by MMDx. The blue dots/ regression line represents no rejection group by MMDx. R indicates spearman’s correlation coefficient with the p-value. The top R in blue indicates no rejection group; Lower R in blue indicates the rejection group. Abbreviations: MMDx: molecular microscopy; dd-cfDNA: donor-derived cell-free DNA.

## Discussion

Collectively, our study is the first to show that dd-cfDNA correlates with both MMDx-based classifications of allograft rejection and tissue RAT profiles of the heart. Cardiac allograft injury and rejection remain one of the most important clinical concerns after heart transplantation (2). Traditional microscopy from EMB is limited by interpretation subjectivity and phenotypic presentation, not reflecting ongoing pathological molecular processes (3, 7). Quantification of dd-cfDNA and the MMDx system both incorporate molecular techniques that allow objective classification and quantification of allograft injury and rejection.

The diagnostic accuracy of dd-cfDNA and MMDx were previously compared with traditional microscopy in multiple clinical studies (4, 7, 8). However, there is a paucity of data assessing the relationship between dd-cfDNA and MMDx. In a recent single-center study, Alam et al. elegantly showed that the concordance across all three diagnostic modalities (dd-cfDNA, MMDx, and traditional microscopy) was quite low (61%), but the comparison between two modalities, the percent agreement was relatively high (70-84%) (10). Similar to their study, our work also shows that the agreement between modalities was substantial (67-75%). In addition, we compared the agreement between modalities based on types of rejection. The percent agreement between dd-cfDNA and EMB-based modalities (MMDx and traditional microscopy) was higher in AMR/ABMR (66-75%), compared to ACR/TCMR (53-54%). This is likely due to the level of dd-cfDNA in the TCMR group not being significantly higher than the no rejection in MMDx. This is also in line with prior studies where higher levels of dd-cfDNA were found in AMR, when compared to ACR (6, 17), and is similar to the relationship between dd-cfDNA and tissue transcript expression in kidney transplantation (11, 12).

A significant gap in current knowledge is the discrepancy between dd-cfDNA levels in ACR and AMR. In this study and in prior work (6), AMR demonstrated significantly higher levels of dd-cfDNA, compared to ACR **(Figure 1A)**. This points to distinct pathogenesis between AMR and ACR leading to differences in the level of dd-cfDNA (6). Based on these data, we posit that AMR results in more vascular injury, which may lead to increased direct release of dd-cfDNA into the circulation. In contrast, ACR is characterized by localized lymphocytic infiltration in the myocardium, which may contain the release of dd-cfDNA. Also of interest is that the non-rejection allograft injury group had elevated levels of dd-cfDNA, potentially owing to the low sample number (N=13).

Molecular microscopy provides mechanistic insight into pathological processes via changes in tissue RAT expression. Therefore, we investigated the relationship between dd-cfDNA, a measure of allograft injury, and tissue RAT assessment by MMDx in an exploratory fashion (7, 9, 17). We demonstrate that dd-cfDNA levels are significantly elevated in RATs classified as abnormal (**Table 2**). This suggests that in allografts expressing abnormal rejection-related transcripts, the circulating allograft injury marker (dd-cfDNA) is also elevated. Although not unexpected, as these RATs collectively constitute the final categorization of allograft rejection and injury by MMDx, it is still of importance to note concordance between distinct and objective diagnostic methods investigating different pathologic pathways.

In addition, there were several RATs as individual transcript or transcript sets that were positively correlated with the level of dd-cfDNA. These results are exploratory in nature and do not confer a causal relationship. However, one transcript of interest was *ROBO4*, the only individual transcript demonstrating a positive correlation with dd-cfDNA. ROBO4 is a member of a roundabout homolog family of neural patterning signals and is expressed selectively in the vascular endothelium (18, 19). ROBO4 interacts with UNC5B, an inhibitor of the VEGF-VEGFR2 signaling pathway, and promotes vascular integrity by counteracting VEGF-dependent endothelial barrier permeability (18, 20, 21). VEGF expression itself is a marker of pro-inflammation and vascular turnover and is associated with allograft rejection and coronary allograft vasculopathy (22). This suggests that the increase in *ROBO4* expression observed in rejection in this study may be compensatory for increased VEGF-dependent signaling in these conditions (23-28). Based on the above, we speculate a common pathway between the release of dd-cfDNA and ROBO4 in the setting of AMR, which may explain the higher level of dd-cfDNA in AMR. Future studies may investigate whether concurrent elevation of both *ROBO4* and dd-cfDNA predicts future rejection. Also, these RNA expression level changes should be confirmed through the immunohistochemistry of ROBO4 protein along with apoptotic and necrotic markers localizing to the endothelium.

There are several limitations to our study. First, the inclusion criteria for this study required the presence of all three diagnostic modalities (dd-cfDNA, molecular microscopy, and traditional microscopy). This may have led to selection bias. Also, we have not differentiated pAMR grade 1I and 1H, which may have differences in the dd-cfDNA level. Second, the time between the dd-cfDNA and EMB varied. However, the date difference was a median of 0 days (interquartile range: 0-4 days). In addition, all dd-cfDNA samples were collected before EMB to prevent transient biopsy-related elevations in dd-cfDNA (29). Third, our study is not designed to evaluate the ability of dd-cfDNA to reduce the need for EMB. Instead, it augments the current knowledge that dd-cfDNA is a reliable noninvasive tool for allograft injury surveillance.

Our work is the first to suggest that combined assessment of dd-cfDNA levels and granular monitoring of tissue RAT may provide synergistic clinical utility in diagnosing rejection. In addition, the broad clustering of related MMDx transcripts into “transcript classifiers” stymies our ability to establish distinct relationships between individual transcripts and dd-cfDNA. An analysis of individual RAT expression patterns within these transcripts may provide even deeper mechanistic insight into rejection and injury, strengthening the combined utility of these tools.

In conclusion, dd-cfDNA levels are elevated in allograft rejection as assessed by molecular microscopy, a finding consistent with previous literature. In addition, based on our exploratory study, we speculate a common pathway between the release of dd-cfDNA and *ROBO4* in the setting of AMR, which may help to explain one mechanism of why dd-cfDNA level is higher in AMR, compared to ACR.

## Data Availability

All data produced in the present study are available upon reasonable request to the authors

## Author Contributions

Study Design: DHL, RW, GHO, BM, PB Data Collection: DHL, AU, VR, TWD Data Analysis: DHL Writing of Manuscript: DHL, RW, TW, RB, VR, TWD, GHO, BM, PB

## Acknowledgments

Part of the study was presented at the 42^nd^ ISHLT Annual Meeting and Scientific Session in 2022 as Oral Presentation. DHL received a Type 1 Travel Award from ISHLT Grants and Awards Committee for this work. The authors would like to thank Renata Ponsirenas (CareDx) for reviewing and editing the manuscript.

